# Endometriosis And Chronic Pelvic Pain Have Similar Impact On Women, But Time To Diagnosis Is Decreasing: An Australian Survey

**DOI:** 10.1101/2020.06.16.20133231

**Authors:** Mike Armour, Justin Sinclair, Cecilia H M Ng, Mikayla S Hyman, Kenny Lawson, Caroline A Smith, Jason Abbott

## Abstract

Chronic pelvic pain (CPP) affects a significant number of women worldwide. Internationally, people with endometriosis report significant negative impact across many areas of their life. We aimed to use an online survey using the EndoCost tool to determine if there was any difference in the impact of CPP in those with vs. those without a confirmed diagnosis of endometriosis, and if there was any change in diagnostic delay since the introduction of clinical guidelines in 2005. 409 responses were received; 340 with a diagnosis of endometriosis and 69 with no diagnosis. People with CPP, regardless of diagnosis, reported moderate to severe dysmenorrhea and non-cyclical pelvic pain. Dyspareunia was also common. Significant negative impact was reported for social, academic, and sexual/romantic relationships in both cohorts. In the endometriosis cohort there was a mean diagnostic delay of eight years, however there was a reduction in both the diagnostic delay (p<0.001) and number of doctors seen before diagnosis (p<0.001) in those presenting more recently. Both endometriosis and CPP have significant negative impact. Whilst there is a decrease in the time to diagnosis, there is an urgent need for improved treatment options and support for women with the disease once the diagnosis is made.

## Introduction

Chronic pain in women and girls has been identified as a top five health concern by Australian women ^1^ and is identified by a government report as an Australian National Women’s Health Priority ^2^. There are a variety of causes for chronic pelvic pain (CPP) including endometriosis, adenomyosis, chronic infection, vulvodynia irritable bowel syndrome or bladder pain syndrome. Endometriosis is the most common cause of CPP ^3^, accounting for 24-40% of all CPP diagnoses ^4,5^ and has a prevalence rate of around 11% in Australia ^6^. Some people have no demonstrated pathology despite detailed surgical investigation or imaging, leading to a diagnosis of Chronic Pelvic Pain Syndrome (CPPS).

It has been repeatedly demonstrated internationally that endometriosis has a long diagnostic delay ^7^, impacts people’s health and wellbeing ^8^, including social activities ^9^, mental and emotional health ^10,11^, work/finances ^9,12^, and sexual relationships ^13^; and those with endometriosis report physical quality of life similar to that of cancer patients ^9^. While most research has focused on endometriosis, it is possible that people with non-endometriosis related CPP experience similar issues but possibly with less severe impact ^14^, however there is a paucity of research in this area.

The aim of this survey was to determine in an Australian population, with endometriosis or non-endometriosis related CPP, the process of diagnosis, the prevalence of different CPP symptoms, and to explore the management and impact on people’s social, sexual/romantic, work, and academic lives.

## Methods

### Questionnaire

The World Endometriosis Research Foundation (WERF) EndoCost tool consists of validated prospective hospital questionnaires and both retrospective and prospective patient questionnaires ^15^. Our study used the 99 item retrospective patient questionnaire, modified to Australian income and ethnicity parameters as per the Australian Bureau of Statistics ^16^. The survey was hosted on SurveyMonkey (www.surveymonkey.com), with an estimated 30-45 minute completion time. The cost of illness burden has been previously published ^12^, with this paper reporting on items contributing to the WERF Global Study on Women’s Health ^9^. It focuses on diagnosis, education, work and social wellbeing as well as recent (3 month) prevalence of other pelvic pain symptoms such as dysmenorrhea and dyspareunia.

### Recruitment

Following ethics approval by the Western Sydney University Human Research Ethics Committee, (approval number H12019), the survey link was distributed via the social media platforms (Facebook, Twitter and Instagram) of Endometriosis Australia, EndoActive and Pelvic Pain Foundation of Australia from February 2017 to April 2017, for a total of eight weeks. The total combined reach of these organizations on social media was just over 35,000 followers at the time of survey distribution. Each organization made two social media posts regarding the survey 3-5 weeks apart. Data collection was closed once there had been no new responses for five days. Informed consent was obtained from all participants. Both the participant information sheet and the survey introduction outlined that completion of the survey implied consent. All research complied with the relevant guidelines and regulations outlined in the National Statement on Ethical Conduct in Human Research (2018)^17^.

### Study population

People were eligible to participate in the survey if they were aged 18-45, currently living in Australia and either had a surgically confirmed diagnosis of endometriosis, or if they had CPP from any cause. CPP was defined as pain in the pelvis for at least six months that caused the person to seek medical attention. Those with CPP either had a laparoscopy that did not show evidence of endometriosis or had not undergone a laparoscopy at the time of survey. This study was designed to measure prevalence of symptoms and assess their impact rather than test a hypothesis, therefore no sample size calculation was performed.

### Analyses

Data were analysed using SPSS v26 (IBM Corporation, Chigago Ill.). Descriptive statistics were presented as means and standard deviations (for normally distributed data), medians and interquartile ranges (for non-normally distributed data), or number and percentages (for categorical data). Inferential statistics for between-group comparisons were performed using a one-way ANOVA, chi-square test or Fishers Exact as appropriate. Correlations between categorical and continuous variables were analysed using Spearman’s rank order correlation, and correlations between two continuous variables were analysed using Pearson’s Correlation. Statistical significance was set at p<0.05. Missing data were reported and not replaced.

To explore the potential effect of the introduction of the first publication of the European Society of Human Reproduction and Embryology (ESHRE) diagnostic guidelines in 2005 ^18^, the 2013 World Endometriosis Society (WES) diagnostic guidelines and updated ESHRE guidelines in 2013 ^19,20^ we planned to group the analysis on diagnostic delay and number of doctors seen prior to diagnosis into three groups based on the timing of their first presentation to a doctor; presentation prior to 2005, presentation between 2005 and 2012, and presentation from 2013 onwards.

## Results

409 valid responses were received. 340 of these people had endometriosis (83%) and 69 (17%) had CPP without a current diagnosis of endometriosis. Table 1 outlines the demographic characteristics of survey participants. The mean age of respondents identifying as being affected by endometriosis was 30.6 (±7) and CPP was 33.7 (±16.3) years. The majority of respondents earned between AUD$510-$1500 per week, with a little more than half of people across both groups held either university or post-secondary qualifications. In people with endometriosis, 33.7% reported stage IV disease at their most recent laparoscopy.

**Table 1:** Demographics.

|  | **Endometriosis**<br>**(n=340)** | **Chronic Pelvic Pain (n=69)** |
| --- | --- | --- |
|  | Mean (SD) | Mean (SD) |
| Age (y) | 30.6 (7.0) | 33.7 (16.3) |
| **Ethnicity, n (%)** |  |  |
| *Caucasian* | 312 (91.8%) | 64 (92.8%) |
| *Asian* | 5 (1.5%) | 2 (2.9%) |
| *Aboriginal/TSI* | 5 (1.5%) | 1 (1.4%) |
| *Polynesian* | 0 (0%) | 0 (0%) |
| *Maori* | 0 (0%) | 0 (0%) |
| *Other* | 17 (5%) | 2 (2.8%) |
| *Blank* | 1 (0.3%) | 0 (0%) |
| **Relationship status, n (%)** |  |  |
| *Single* | 69 (20.3%) | 14 (20.3%) |
| *Married / Defacto* | 211 (62.1%) | 50 (72.5%) |
| *In a relationship but not living with partner* | 49 (14.4%) | 4 (5.8) |
| *Divorced / Separated* | 9 (2.6%) | 0 (0%) |
| *Widowed* | 0 (0%) | 1 (1.4%) |
| *Blank* | 2 (0.6%) | 0 (0%) |
| **Occupation, n (%)** |  |  |
| *Self-employed* | 23 (6.8%) | 4 (5.8%) |
| *Employed* | 236 (69.4%) | 47 (68.1%) |
| *Going to school or University* | 70 (20.6%) | 15 (21.7%) |
| *Home duties/ caring for children and family* | 43 (12.6%) | 10 (14.5%) |
| *Doing voluntary work* | 18 (5.3%) | 2 (2.9%) |
| *Unable to work due to pelvic pain symptoms* | 23 (6.8%) | 7 (10.1%) |
| *Unable to work for other reasons* | 5 (1.5%) | 2 (2.9%) |
| **Level of education, n (%)** |  |  |
| *Primary School* | 0 (0%) | 0 (0%) |
| *Lower secondary* | 24 (7%) | 2 (2.9%) |
| *Upper secondary* | 47 (13.8%) | 6 (8.7%) |
| *Post-secondary (TAFE)* | 91 (26.8%) | 22 (31.9%) |
| *University* | 123 (36.2%) | 25 (36.2%) |
| *Post graduate* | 55 (16.2%) | 14 (20.3%) |
| *Blank* | 0 (0%) | 0 (0%) |
| **Currently have children, n (%)** |  |  |
| *Yes* | 97 (28.5%) | 23 (33.3%) |
| *No* | 242 (71.2%) | 46 (66.7%) |
| *Blank* | 1 (>1%) | 0 (0%) |
| **Endometriosis stage at most recent laparoscopy, n (%)** |  |  |
| *Stage 1* | 14 (4.1%) |  |
| *Stage 2* | 51 (15%) |  |
| *Stage 3* | 75 (22.1%) |  |
| *Stage 4* | 115 (33.7%) |  |
| *Can't remember* | 57 (16.8%) |  |
| *Blank* | 28 (8.3%) |  |

### Symptoms and diagnosis

Table 2 outlines the diagnostic data and symptoms at onset for both cohorts. A Pearson’s correlation found a statistically significant, strong negative correlation between the year when medical attention was first sought and the number of years taken to get a diagnosis of endometriosis, *r*=-0.71, p<0.001. We compared the delay between when medical attention was first sought and found a significant difference in delay for the person who first sought medical attention prior to 2005 (9.9±6.6 years), between 2005 and 2012 (4.8±2.6 years) and from 2013 (1.5±0.7 years) onwards (p<0.001). A Spearman’s correlation was performed and found there was a statistically significant negative correlation between the year when medical attention was first sought and the number of doctors seen to get a diagnosis in people with endometriosis, r_s_=-0.305, p<0.001. Similar to the reduction in delay, the number of doctors seen before diagnosis was also reduced significantly after the introduction of the first ESHRE guideline (p<0.001).

**Table 2:** Symptoms and diagnosis.

|  | **Endometriosis**<br>**(n=340)**<br>**Mean (SD)** | **Chronic Pelvic Pain**<br>**(n=69)**<br>**Mean (SD)** | **P value** |
| --- | --- | --- | --- |
| **Age when symptoms first appeared** | 18 (6) | 21.9 (8.6) | <0.001* |
| **Time between symptom onset and first doctors visit (years)** | 2.9 (4.4) | 1.8 (3.4) | 0.07 |
| **Time between doctors visit and diagnosis of endometriosis (years)** | 4.9 (5.7) | N/A | N/A |
| **Number of Doctors seen before diagnosis of endometriosis** | 5 (4) | N/A | N/A |
| **Number of doctors seen by year of first presentation** |  |  |  |
| *Prior to 2005* | 7.3 (7.0) | N/A | N/A |
| *Between 2005-2012* | 4.3 (3.2) | N/A | N/A |
| *2013 onwards* | 3.9 (2.6) | N/A | N/A |
| **Time between year of first presentation and diagnosis** |  |  |  |
| *First sought medical attention before 2005* | 9.9 (6.6) | N/A | N/A |
| *First sought medical attention between 2005-2012* | 4.8 (2.6) | N/A | N/A |
| *First sought medical attention after 2013* | 1.5 (0.7) | N/A | N/A |
| **Symptoms at onset, n (%)** |  |  |  |
| Severe Dysmenorrhea | 260 (89.4%) | 57 (82.6%) | 0.66 |
| Deep Dyspareunia | 84 (32.3%) | 27 (39.1%) |  |
| Pelvic Pain | 229 (78.7%) | 51 (73.9%) |  |
| Ovulation pain | 136 (46.7%) | 29 (42%) |  |
| Cyclical or peri-menstrual symptoms (e.g. bowel or bladder associated)<br>with or without abnormal bleeding | 111 (38.1%) | 33(47.8%) |  |
| Infertility | 22 (7.6%) | 6 (8.7%) |  |
| Chronic Fatigue | 113 (38.8%) | 25 (36.2%) |  |
| Blank | 25 (7.4%) | 0 (0%) |  |

\*Represents significance (p<0.001)

There was a moderate negative correlation between the year symptoms first appeared and the delay in seeking medical attention, *r*=-0.39, p<0.001.

People experiencing dysmenorrhea with every period in the previous three months were comparable between the endometriosis (85.9%) and CPP (87.0%) cohorts, and dyspareunia was common in both groups, with over two thirds of respondents with endometriosis (69.1%) and CPP (66.7%) reporting experiencing pelvic pain either with or in the 24 hours after intercourse (Table 3).

**Table 3:**
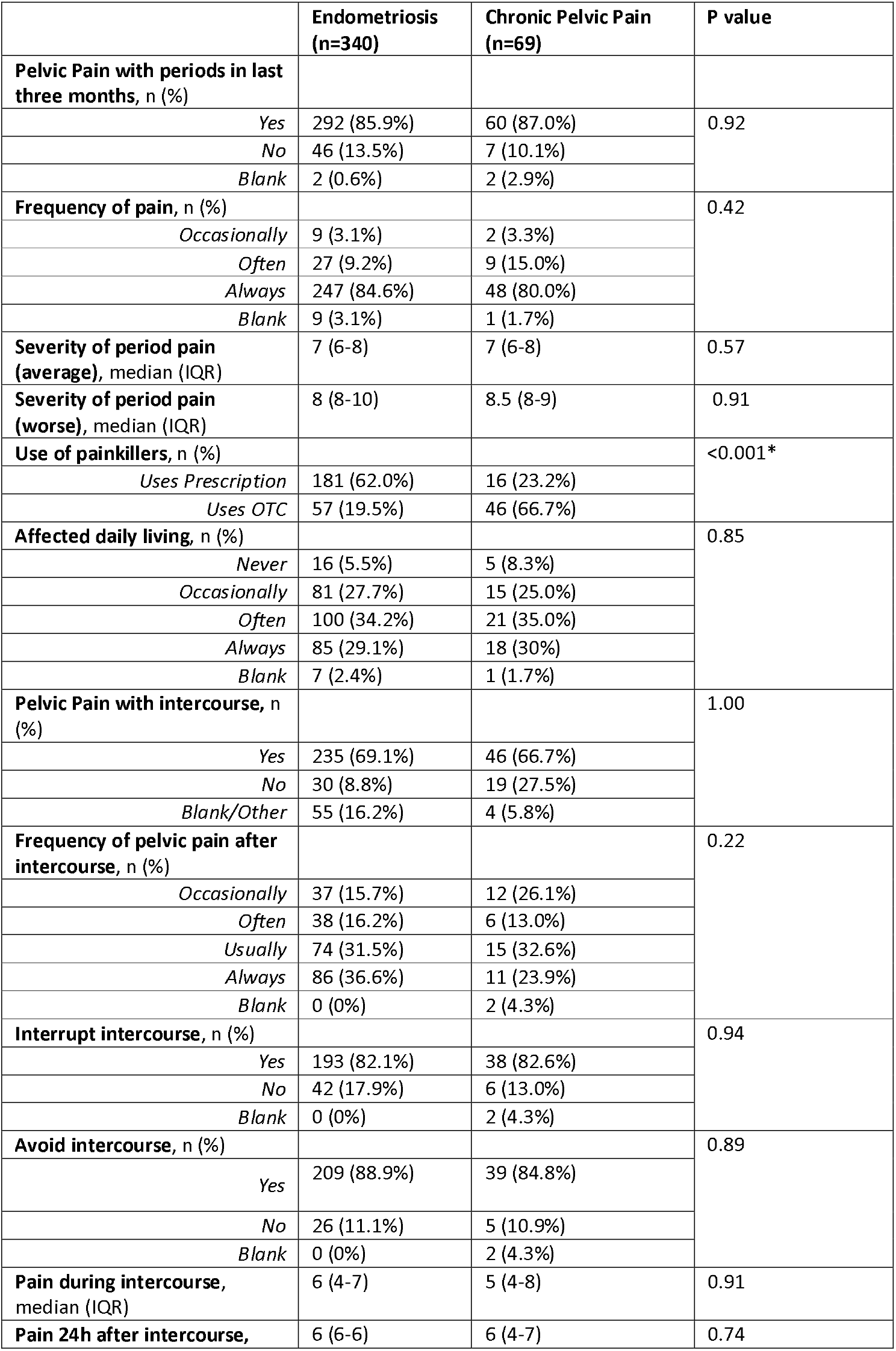
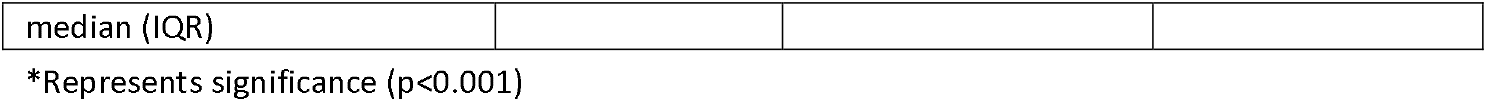
Description of menstrual symptoms and sexual intercourse.

### Social, Educational and Work impact

Substantial negative impact was reported across both cohorts in education, employment and social domains (Table 4).

**Table 4:**
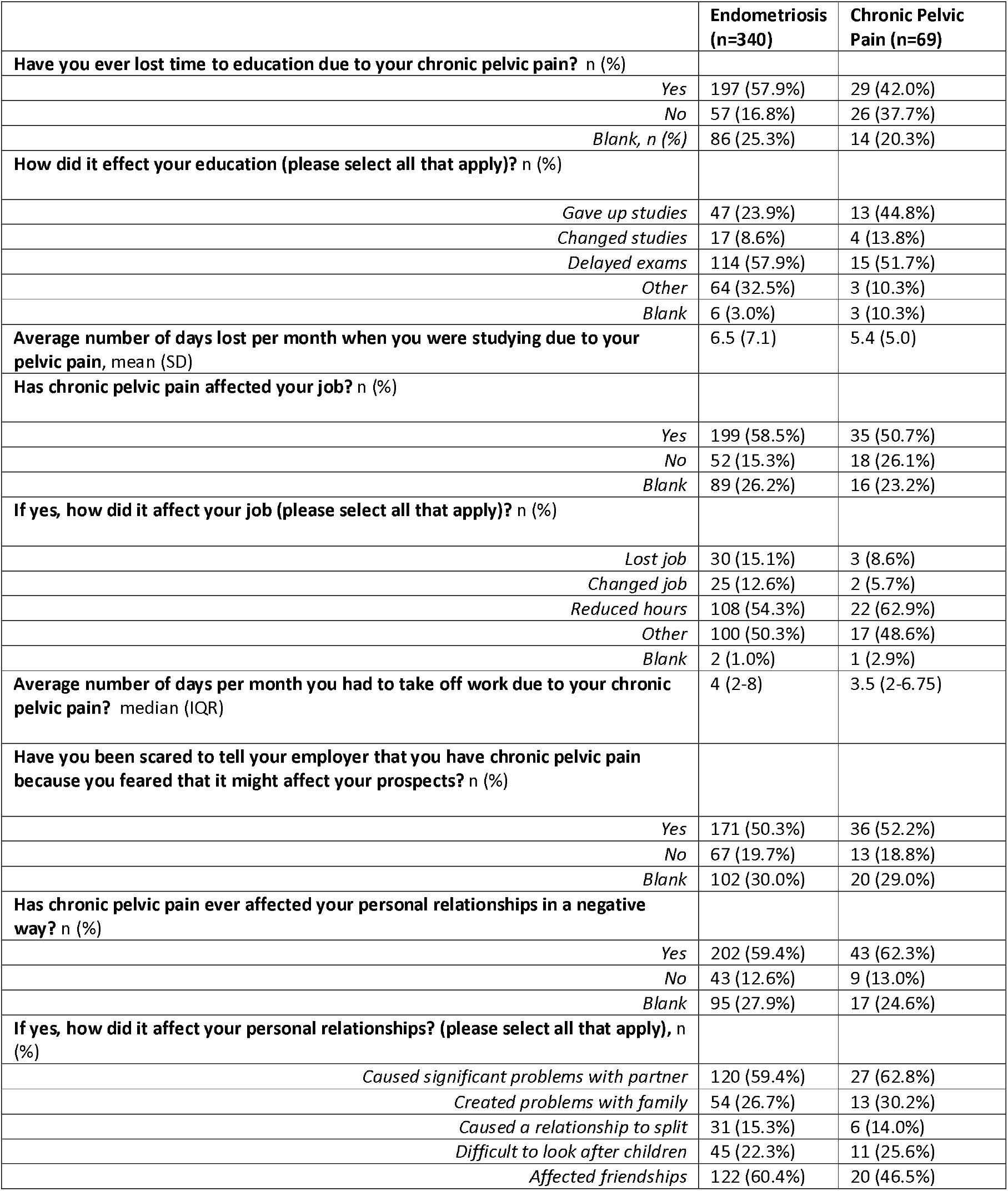
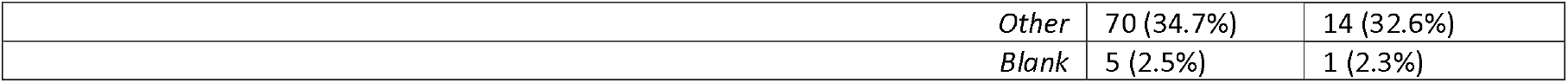
Social, Educational and Work impact.

Personal relationships were similarly affected between cohorts, with more than half of respondents indicating problems with their sexual or romantic partners. For those who said pelvic pain impacted their personal relationships around one in six people in either group indicated that this had caused the breakdown of their relationship. The most commonly reported impact for people with endometriosis (35.9%) was that it affected their friendships. At least one in five people in each cohort indicated that they found it difficult to look after children, and almost a third indicated it caused issues with their family.

### Non-cyclical pelvic pain

Supplementary Table S1 compares the prevalence, severity and management of non-cyclical pelvic pain between cohorts.

## Discussion

Data from this study demonstrates that people with CPP, irrespective of diagnosis, commonly report moderate to severe pelvic pain symptoms, consistent with recent research on endometriosis ^21^. People with endometriosis frequently report symptom onset (most commonly dysmenorrhea and pelvic pain) in their late teens and overall report an eight year diagnostic delay from symptom onset. Nearly three years of this delay is due to the person not presenting to their medical care provider and the remaining 5 years between initial presentation to a doctor and a surgical diagnosis of endometriosis. This is similar to the delay reported in other studies ^9,22^ and may, at least be in part, be due to a normalization of the severe dysmenorrhea that is frequently reported in young people with pelvic pain ^23-25^. It may also represent commonly held beliefs that even moderate or greater severity menstrual or pelvic pain is a normal part of ‘becoming a woman’ or ‘something you need to put up with’ as documented in qualitative studies ^22,26,27^. However, our findings indicate that this delay in presentation to a medical professional is reducing over time, with those reporting their symptoms that have started more recently showing a shorter time delay in seeking medical attention. This may be due to an increased awareness of the symptoms of endometriosis and encouragement to seek medical assistance for these through the efforts of advocacy organizations and the recent increased media coverage of endometriosis including events such as Endometriosis Awareness Month. A variety of educational programs, both in person and online, designed to improve awareness of menstruation and endometriosis have also been developed for Australia and elsewhere ^28-32^. Most of these resources have been implemented in 2019 and beyond, therefore, given the timing of our survey, it is too soon to see whether these programs may contribute to any further reduction in diagnostic delay.

Similarly, the clinician seeing the person with CPP for the first time may normalize those symptoms ^22^ or provide other diagnoses such as irritable bowel syndrome or sexually transmitted infections ^27,33^, which can lead to frustration for the patient and further delay in appropriate management that targets their symptoms. Since the introduction of the ESHRE guideline on endometriosis in 2005, there appears to be a significant reduction in both the number of doctors seen before diagnosis and the time between seeking medical help and a diagnosis of endometriosis. It is unclear at this time if an earlier diagnosis of endometriosis will improve pain and fertility outcomes. However, earlier identification of abnormal menstrual symptoms in early education programs ^28^ may allow for an earlier intervention that could change the course of disease and, at least, improve the quality of life of those affected if appropriately managed. This goal is identified in the Australian National Action Plan for Endometriosis ^34^ and needs further investigation in future research studies.

Pain and the associated symptom of fatigue is commonly seen in people with endometriosis ^21,35^ and are significant contributors to both the work and education-related absenteeism and presenteeism seen in our respondents ^27^. Fatigue is a common symptom in people with endometriosis and other types of pelvic pain with 39-43% of respondents reporting that symptom at the onset and all clinicians should ask about this symptom when taking a history of a person presenting with pelvic pain. The fatigue experienced may be debilitating ^36^ and there is a strong correlation between pain severity and fatigue ^37^, almost certainly explaining why chronic fatigue syndrome is much more common in people with endometriosis than the general population ^37^. Educational absenteeism in our survey led to giving up study for 25-50% of people. This is notable since academic achievement declines with any absenteeism ^38,39^. Absenteeism also increases social isolation and decreases engagement with peers. The community ^40^ and the consequential cessation of education may substantially limit long-term opportunities for people with endometriosis and CPP.

Previous international surveys ^8,9^ and qualitative research ^22,27^ have highlighted the impact of endometriosis on social activities and marital/sexual relationships. We also demonstrated that personal relationships, whether friendships or those of a romantic/sexual nature, were negatively impacted by endometriosis and CPP. Reasons for this may include the fatigue experienced reducing social interactions ^35,37^ and increased anxiety of experiencing heavy menstrual bleeding, nausea, gastrointestinal and bladder symptoms ^27^ in public places when interacting with friends, family or others. Similar to previous work ^13^, romantic and sexual relationships were commonly affected in both cohorts, with pain as a direct contributor, but fatigue, nausea, depression, and other co-morbidities also, often, contributing to relationship strain and potential breakdown ^41^.

Compared to those with endometriosis people with CPP had symptom onset later, in their early 20s, but with similar presenting symptoms. Previous work also identified a substantial number of people who report CPP symptoms but have not been given any diagnosis ^42,43^. Of course, pelvic pain may later be diagnosed as endometriosis ^21^ but most importantly, our work confirms that both diagnosed and non-diagnosed CPP causes substantive negative impact on work, education, sexual relationships, family and friendships. It is therefore necessary to identify and have a treatment plan for pelvic pain symptoms, regardless of the current or future diagnosis.

There are a number of strengths of this study. Whilst this is the first Australian study for endometriosis and there exist similar estimates from other countries ^8,44,45^, there are no previous data on the impact of CPP. The use of the WERF EndoCost tool also allows comparison between datasets generated between different centers. However, there are limitations of this study that must be acknowledged. The overall sample size is small, which may reflect the in-depth nature of the questionnaires, which required up to 45 minutes to complete. Respondents were asked to answer the endometriosis section only if they had a histologically confirmed surgical diagnosis although confirmatory evidence of that was not possible. We recognize that people designated to the CPP cohort may have undiagnosed endometriosis, which is an eventual diagnosis in 33-75% ^36,37^ of this group. Importantly, our study demonstrates similar negative impacts across many domains irrespective of diagnosis. We did not collect data on geographical location of respondents, so it is possible that our sample did not represent potential variations in the access to appropriately specialised health care professionals, work, or educational opportunities. Respondents were predominantly Caucasian, and therefore caution must be heeded in extrapolating these findings to other culturally and linguistically diverse groups. Given the timing of the survey in early 2017, there may be a ceiling effect with respect to diagnostic delay for the cohort that presented to their health care professional after 2013. However, the reported diagnostic delay observed between 2005-2012 is unlikely to be affected by any ceiling effect. Finally, our recruitment was almost solely via advisory and support groups on social media, and people recruited via this method are likely to have more severe symptoms than those recruited in other settings such as tertiary care ^46^. So it is possible that our respondents may be biased towards a more severe impact of their symptoms on their lives and may not represent the population of people with endometriosis as a whole.

## Conclusion

People with CPP, regardless of a diagnosis of endometriosis, experience significant negative impact across a range of domains including education, work, social, and romantic/sexual relationships, which may have long lasting consequences if not appropriately recognised and managed through access to adequate health care resources. Our findings suggest that there has been a significant decrease in diagnostic delay over time, possibly due to improved guidelines and advocacy. However, the diagnostic delay still exists and still carries a significant burden to those suffering CPP. Widespread, evidence based educational programs, recognising the symptom profile of CPP, for both the public and medical providers, are urgently needed to help reduce this delay and implement and provide access to the most appropriate management plan for these people.

## Data Availability

The datasets generated during and/or analysed during the current study are available from the corresponding author on reasonable request.

## Acknowledgements

Thank you to the organisations that promoted this survey; Endometriosis Australia, EndoActive and the Pelvic Pain Foundation of Australia. Thank you to the World Endometriosis Research Foundation (WERF) for allowing us to use their EndoCost tool and to Lone Hummelshoj who provided valuable guidance on the final manuscript.

## Author contributions

All authors contributed significantly to this work. MA, KL, CS and JA designed the study. MA, KL, JS and MH performed the data collection and statistical analysis. MA and JS drafted the article. JA, CS, CN and KL contributed to critical revisions on the manuscript. All authors reviewed the manuscript and approved the final draft.

## Competing interests

MA,JS, CS: As a medical research institute, NICM Health Research Institute receives research grants and donations from foundations, universities, government agencies and industry. Sponsors and donors provide untied and tied funding for work to advance the vision and mission of the Institute. This study was not specifically supported by donor or sponsor funding to NICM. MA is a member of the clinical advisory board for Endometriosis Australia.

JA: Medical Director, Endometriosis Australia (NFP)

KL: None known.

CN: None known.

MH: None known.

## Notes

### Funding Statement

No external funding recieved

### Author Declarations

Western Sydney University Human Research Ethics Committee, (approval number H12019)

